# Segmental Lung Sound Analysis in Obstructive Lung Diseases Using Electronic Stethoscope; a protocol to establish an acoustic repository

**DOI:** 10.64898/2026.05.27.26354263

**Authors:** RPH Anuradha, D Yasaratne, GMRI Godaliyadda, MP Ekanayake, R Severin

## Abstract

**Introduction:** Obstructive lung diseases (OLDs) are responsible for high rates of illness and death worldwide. Inflammation, chronic airflow limitation, and bronchial remodeling occur in OLD and eventually result in the unique respiratory sounds. Despite its subjective and having low reproducibility, still traditional auscultation using a manual stethoscope is the main method used to identify the lung sounds. Nevertheless, the combination of recent advancements in digital stethoscopes and AI (Artificial Intelligence) has permitted the objective measurement of lung sounds. Nevertheless, there is a lack of standardized, region-specific databases for AI training and validation. Even though lung sound classification is an emerging aspect in research and telerehabilitation the lobar wise acoustic pattern is still novel due to lack of prevailing database to train AI models. Identifying this gap this study aims to develop an acoustic repository and analyze the data using segmental lung sounds from patients with OLDs and healthy controls through an electronic stethoscope.

**Methods and analysis:** This is a cross sectional observational study involving 120 participants (60 OLD patients and 60 healthy controls). Lobar wise acoustic signals will be captured using an electronic stethoscope in healthy and diseases population. The data will be analyzed using Audacity software for annotations and then it will be used for feature extraction and statistical analysis. The acoustic features extracted through Audacity, will include frequency, intensity, pitch, and root mean square (RMS) energy. Repeated measures ANOVA will be applied to compare mean sound intensities across lung segments while Pearson correlation will be used to assess associations with body composition parameters. The data will then be standardized for AI-based diagnostic applications.

**Ethics and dissemination:** The study is being reviewed from the Ethics Review Committee, Faculty of Medicine, University of Peradeniya (2025/EC/87) will be sought. Informed consent will be obtained in writing. The dissemination of results will take place through peer-reviewed publications and the creation of a public database containing lung sounds from the region.

## 1. Introduction

Obstructive Lung Diseases (OLD) are a group of lung diseases of the respiratory tract that involve persistent obstruction of bronchial airflow, often associated with chronic inflammation of airway epithelium, bronchi, and bronchioles(1), which cause increased airway resistance, inflammation and changes in the structure of the respiratory system(2). Chronic inflammation in OLD leads to persistent airflow limitation, increased airway resistance, and structural changes in the respiratory system(2). Common types of OLD include Chronic Obstructive Pulmonary Diseases (COPD), Asthma, Bronchiectasis, and Cystic Fibrosis (CF). While chronic bronchitis is marked by increased inflammation of bronchial tubes alongside a productive cough, emphysema is the stage that leads to the destruction of the walls of the alveoli(3).The occurrence of an obstructive type of respiratory disease is typically chronic and progressive, whereas asthma is reversible with treatments(4). OLDs account for symptoms such as dyspnea, wheezing, and reduced exercise tolerance(5). In this type of disease, hyperinflation of the lungs is a common phenomenon, where mucus plugging and prolonged bronchoconstriction oppose full deflation of the lungs. This generates a feeling of air hunger in a patient, particularly during exertion or at night(6). Diminished breath sounds and limited air entry are characteristic features of OLD. This reduction of breath sounds occurs due to multiple mechanisms: airway constriction, loss of alveolar elasticity, and dynamic hyperinflation of the lungs. Air trapping due to mucus plugging and bronchoconstriction further causes non-uniform airflow distribution and reduces the intensity of lung sounds(7).

Auscultation provides real-time assessment of airflow and helps identify abnormal sounds such as wheezes, rhonchi, and crackles. In OLDs, diminished breath sounds reflect airway narrowing and air trapping(7). Yet, manual auscultation suffers from limitations—low reproducibility, inter-observer variability, and inability to quantify sound intensity. Furthermore, segmental differences in breath sounds are often neglected, though these may provide information into disease localization(8,9).

Muscle mass also makes a significant contribution to the conduction of breath sounds. While added muscle density in athletes may help in improved sound conduction, overdeveloped musculature may dampen breath sounds, particularly in the posterior thoracic regions(10). Research findings show that age and sex have major influences on the characteristics of lung sounds(11). It is discovered that male possesses lower-pitched breath sounds due to greater lung volumes and longer airway lengths. Females and older subjects possess higher-pitched lung sounds that can affect clinical auscultation findings(12). Thoracic compliance and elastic properties of the lungs also affect the variations in breath sound and amplitude ^11^.

Lung Auscultation using a stethoscope can be considered a simple, non-invasive, and real-time approach to assessing the respiratory condition of OLDs. The simplicity and portability of a stethoscope make it a valuable instrument in clinical practice, primary care, and remote medical consultations, where the assessment of lung function is carried out quickly and directly without the need to refer to high-technological devices(13). Unlike diagnostic investigations such as spirometry or computed tomography (CT) scans that require specialized settings and equipment, stethoscope auscultation can be done easily at the bedside, in the outpatient department, and in emergency medicine and hence is a valuable initial screening test (14).

Machine Learning and Artificial Intelligent-based identification techniques have greatly enhanced breath sound analysis with the help of large datasets and deep learning algorithms to detect different respiratory sounds with high accuracy. In addition, computerized breath sound analysis, such as Mel-frequency cepstral coefficients (MFCCs), have proven useful in detecting wheezing and crackles(15).

The digitally recorded sounds were subsequently interpreted by expert clinicians, often blinded to the initial clinical findings, in an attempt to minimize observational bias (16). The literature showed high concordance rates between digital and traditional auscultation (17). Littman digital stethoscopes highlighted its potential advantages, including amplification of low sounds, noise cancellation effect. Consequently, Littman digital stethoscopes are now regarded as valuable adjuncts to the traditional clinical examination, enhancing diagnostic accuracy and promoting greater access to specialist services (18).

This study aims to develop a standardized, segmental digital lung sound repository of OLD patients using a validated electronic stethoscope. The resulting dataset will serve as the foundation for future AI-based diagnostic algorithms, enhance telemedicine applications, and improve clinical decision-making in low-resource settings.

## 2. Project Objectives

### 2.1 Primary Objectives

1. Characterize segmental and lobar abnormalities in lung sound characteristics among patients with OLDs compared with healthy controls.
2. Quantify Root Mean Square (RMS) intensity, dominant frequency, and spectral distribution in standardized anatomical lung regions.
3. Develop an electronic repository of lung sounds that will include annotated sound files with corresponding clinical metadata for both OLD patients and healthy participants.
4. Data will serve as a reference framework for future training and validation of AI-based respiratory diagnostic tools.
5. Establish benchmark parameters for segmental breath sound recording applicable to both clinical and telemedicine applications.

### 2.2 Secondary Objectives

1. Establish a publicly accessible, repository of anonymized lung sound recordings and metadata for future research collaboration and educational use.
2. Identification of distinct segmental acoustic signatures in obstructive lung disease. Establishment of a replicable and open-source methodology for clinical and research-based digital auscultation.

## 3. Study Population

This study is a prospective, cross-sectional study. It will compare segmental respiratory sound characteristics between OLD patients and healthy controls and serves as a data repository for AI based model training for identifying lobe wise respiratory patterns. This study will be conducted in the respiratory clinic at teaching hospital, Peradeniya and the respiratory clinic at the faculty of Medicine, University of Peradeniya. All recordings will occur in sound-controlled environments to minimize background interference. Patients will be recruited using the following inclusion criteria. Age- and sex-matched healthy non-smokers recruited from university and community populations.

Inclusion Criteria

- Clinically diagnosed OLD
- Age between 40 and 75 years.
- Stable condition without exacerbation in the previous month.
- Ability to follow breathing instructions.

Exclusion criteria

- Acute respiratory infection or recent thoracic surgery (<6 months).
- Severe cardiovascular or neurological comorbidities.
- Cognitive impairment or refusal to consent.

### 3.1 Sample size calculation

Confidence Interval is assigned as 95%

α is assigned as 0.05, Z - 1.96

Standard deviation: σ = 1 (conservative sample assumption) d = 0.3 (margin of error for exploratory descriptive studies) n = required sample size

E = 0.5 (acceptable margin of error)

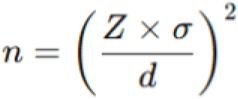

43 participants will be assigned per group. Adding 15% drop out rates, 43+15%≈50.

Therefore 50 participants will be taken for the OLD sample group and 50 participants will be taken for the healthy group.

## 4. Methodology

Demographic (age, sex, BMI, smoking status) and clinical variables (disease duration, medications, comorbidities) will be recorded from the medical reports of the patients and interview based data collection will be conducted for the healthy population. Baseline vitals (heart rate, respiratory rate, BP, SpO₂) will be measured.

A digital stethoscope will be used as the primary device for recording respiratory sounds during segmental breathing exercises. Unlike traditional stethoscopes, electronic stethoscopes offer amplification, real-time recording, noise filtering, and digital storage, which are extremely suitable for acoustic data analysis and further integration with machine learning. The device used here will be Littmann 8480.

To maintain consistency and accuracy in data collection, participants will be positioned comfortably in sitting, supine, and side lying postures during auscultation. For participants unable to maintain an upright posture due to health constraints, necessary adjustments will be made to accommodate their comfort while ensuring that data quality is preserved.

Standardized anatomical landmarks will be taken as the auscultation points of the subjects.

- 2nd intercostal space, midclavicular line (anterior)
- 4th–5th intercostal space, midaxillary line (lateral)
- 7th intercostal space, scapular line (posterior)

Participants perform three breathing cycles per point, with 10-second rest intervals. Audio captured in .WAV format using a Littman 8480 digital stethoscope. Recordings of one lobe will last 10–15 seconds per participant.

### 4.1 Status of the study

At the time of manuscript submission, participant recruitment and data collection has not started yet. Recruitment and data collection will commence in June 2026 at Teaching hospital, Peradeniya and at the university of Peradeniya, Sri Lanka. Data collection expected to continue until May 2027. Data collection is expected to be completed by June 2027.

Preliminary pilot recordings and technical validation procedures have been performed to optimize the electronic stethoscope reliability with a single subject; however, no formal outcome analysis related to the study objectives has been conducted at the time of submission.

The study has not yet generated results, and statistical analysis will commence only after completion of data collection. Final study findings are expected to be available by August 2027.

## 5. Data Management and Data Protection

Audio files containing multiple breath cycles will be imported into Audacity, a free open-source audio editing software. Audacity 3.6 can be used as a lung sound processing software as it poses high resolution waveform visualization and labelling capacity. Preprocessing will be done using Audacity 3.6, involving high-pass and low-pass filtering to remove background noise and artifacts. Each breathing segment will be visualized using waveform analysis and manual annotations will be taken place for the auditory characteristics of the waveform. The inspiration and expirations phases of each respiratory cycle will be segmented and labeled for feature extraction. Using Audacity’s “Add Label at Selection” function, the onset and offset of breath cycles will be marked, and the audio will be cut into labeled clips via the “Export Multiple” feature. These clips, exported as .wav files, will be sorted by lung segment and condition.

Acoustic feature extraction and signal processing will be done by using validated computational audio analysis libraries in Python (Librosa) to ensure AI based pipelines for data and R studio soft ware to ensure methodological statistical analysis.

**Figure 1:**
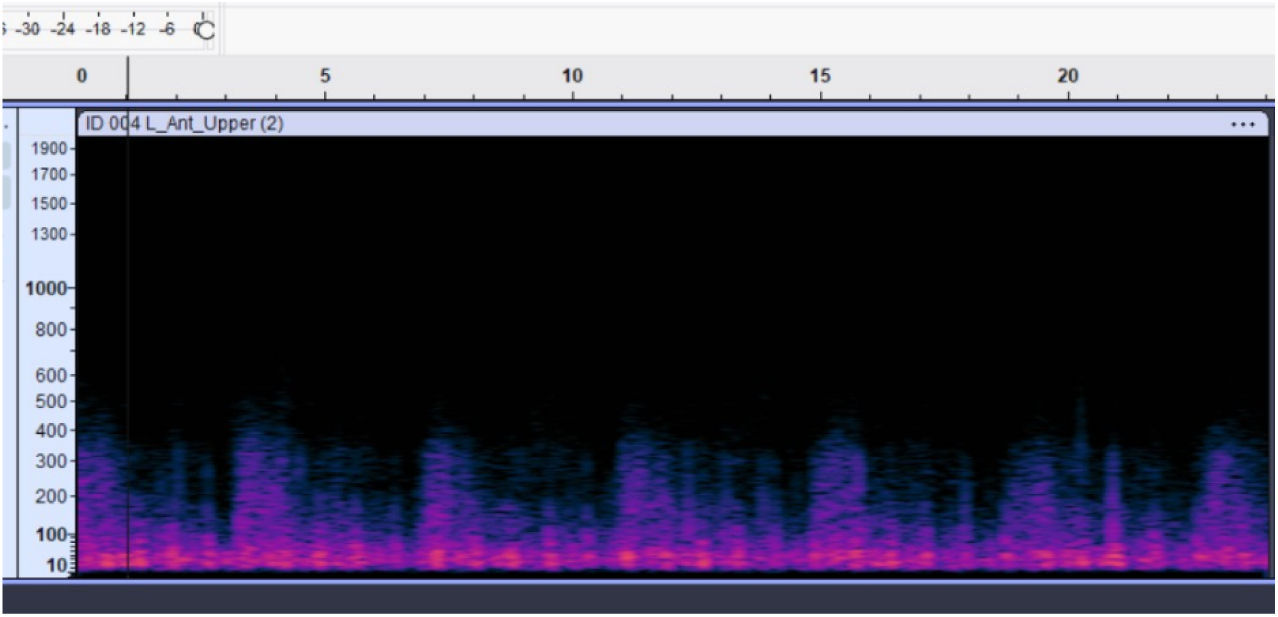
Representative waveform in spectrogram waveforms using Audacity Soft wear.

Thus, each respiratory cycle will be annotated for its inspiratory and expiratory phases. Added sounds can be annotated manually for wheezing and rhonchi. The annotation will be done by a respiratory therapist and will be confirmed by a consultant respiratory physician. Spectrograms will generate time-frequency representations, allowing detection of pathological features such as crackles and wheezes with the aeration for specific lung segments. Combining spectrogram and waveform analyses enhances precise classification of lung sound abnormalities.

Lung sound recordings will be annotated to mark inspiratory and expiratory phases in order to train artificial intelligence data models. Key acoustic parameters like amplitude, frequency, and signal intensity will be extracted and exported numerically for statistical analysis. To ensure methodological validity, a subset of recordings will undergo parallel feature extraction using both Audacity-assisted preprocessing and Python-based Librosa analysis.

### 5.1 Access to Data

The accessibility of the final dataset and lung sound repository will be managed according to strict data ethics and protocols. The primary investigator and selected research members at University of Peradeniya will be primarily entrusted with all data management. The lung sound repository will be subjected to strict protocols to ensure confidentiality.

In line with the principles of open science and collaboration in clinical research, the completed and anonymized digital library of lung sounds will be provided to authorized clinicians, respiratory physiotherapists, and researchers from recognized institutions. Sharing will be carried out after careful review and approval from the Ethics Review Committee, Faculty of Medicine, University of Peradeniya, to ensure that it keeps within the originally intended framework and purposes.

**Figure 2:**
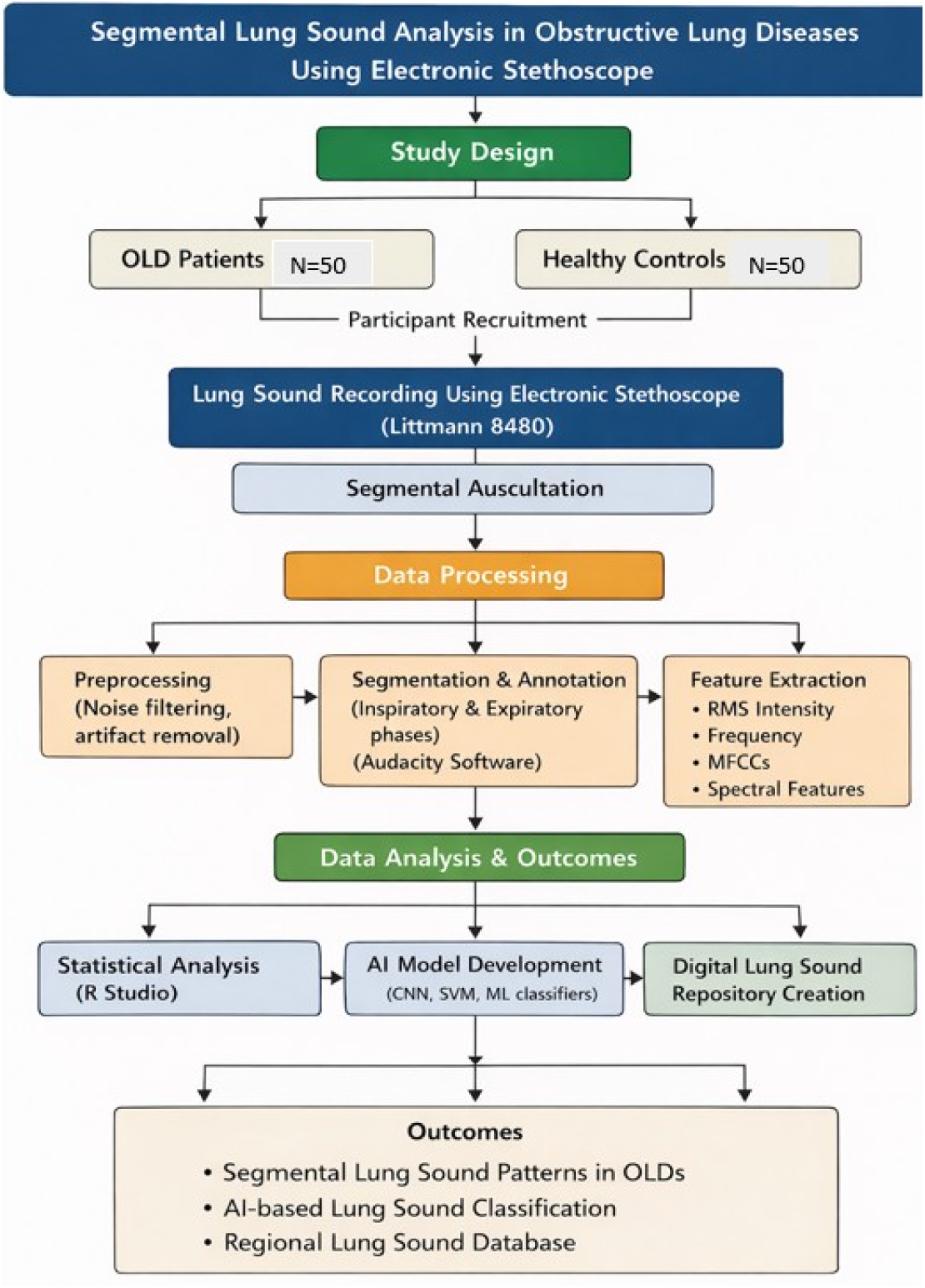
Workflow of the study.

**Table 1:**
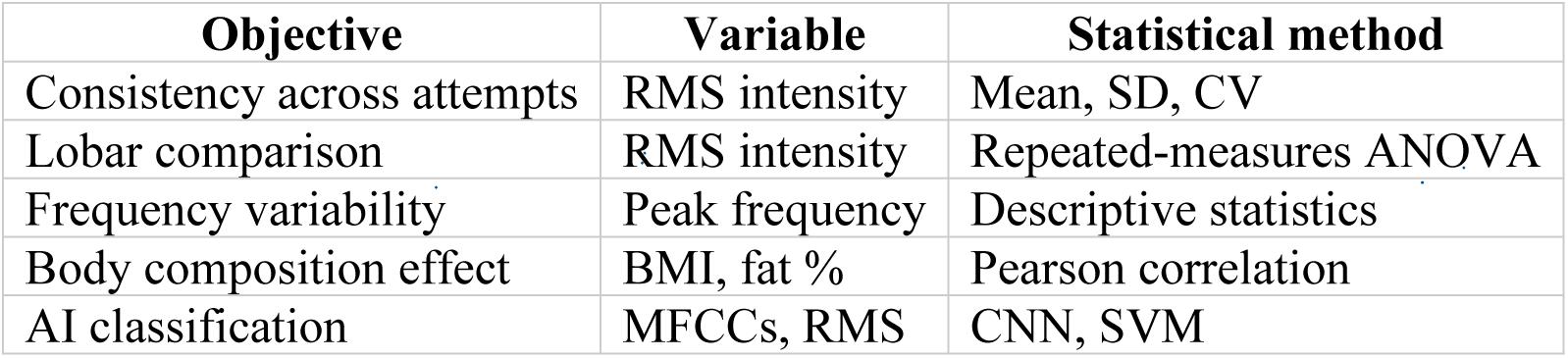
Planned statistical analysis.

| Objective | Variable | Statistical method |
| --- | --- | --- |
| Consistency across attempts | RMS intensity | Mean, SD, CV |
| Lobar comparison | RMS intensity | Repeated-measures ANOVA |
| Frequency variability | Peak frequency | Descriptive statistics |
| Body composition effect | BMI, fat % | Pearson correlation |
| AI classification | MFCCs, RMS | CNN, SVM |

## 6. Discussion

The main objective / intention of this study is the quantitative analysis of segmental and lobar variations in lung sound characteristics among patients with clinically confirmed obstructive lung diseases and healthy controls. The analysis will compare intensity, frequency spectra, and waveform morphology in the six major anatomical lung regions in both anterior and posterior aspects: Right Upper, Right Middle, Right Lower, Left Upper, and Left Middle and Left Lower.

The primary analysis will involve the calculation of RMS intensity in dB and dominant frequency in Hz of the recorded lung sounds. RMS intensity will represent the amplitude or energy of the sound signal, which corresponds to the loudness of breath sounds. Frequency domain analysis using methods such as Fast Fourier Transform (FFT) and Mel-Frequency Cepstral Coefficients (MFCCs) is proposed to detect spectral variations and pathological acoustic patterns such as wheezes, crackles, and diminished air entry.

Intra-individual variability and differences between groups for lobar RMS intensity and frequency will be compared. Using this approach, regional ventilation heterogeneity will be able to be determined.

For each subject, mean RMS values and mean dominant frequencies will be determined for each lung lobes. The results will be normalized based on background noise levels and body composition measures (BMI, percentage of body fat) to reduce possible confounding factors in sound wave conduction. To identify differences among lobes, repeated-measures ANOVA tests will be conducted, while Pearson correlation tests will be used to explore associations between pulmonary sound properties and body measurements, as well as pulmonary function test results.

The reason for choosing primary outcome measurement in this manner is because obstructive pathologies generally follow a heterogeneous pattern of airflow limitation to produce differences in intensity and frequency of breath sounds in different areas because of air trapping, mucus plugs, and segmental collapse. These differences will then enable an objective measurement of the intensity of the disease, which could potentially improve the diagnostic yield of auscultation.

The secondary endpoints include the establishment of an electronic database of respiratory sounds and the use of artificial intelligence models for disease prediction through automated classification.

The entire lung sounds recording data collected in the course of the project will form the basis for a digital archive consisting of identified sound files together with demographic-physiological metadata. The sound files will be categorized according to participant group (obstructive lung disease patients or healthy controls), anatomical lung region (right upper, right middle, right lower, left upper, and left lower lobes), chest surface location (anterior and posterior), and respiratory phase (inspiration and expiration).

This online repository will help formulate a national database. This database will help train researchers on respiratory sounds. This online repository will only be accessed through an encrypted channel on an institutional server. This online repository will contain research-standard open-access data. This will help people access this research data free of cost.

A subset of this carefully crafted dataset will be used to train supervised machine learning algorithms (such as convolutional neural networks) for automatic identification and classification of lung sounds. These algorithms will be trained on spectrograms and feature vectors computed using MFCCs, RMS energy, zero-crossing rate, and spectral centroid.

The proposed model’s performance is assessed using accuracy, sensitivity, specificity, and F1-score methods, which are then validated on the data set using 10-fold cross-validation. The objective is to develop an AI-assisted auscultation system for distinguishing between normal patterns, obstructive patterns, segmental patterns, and pinpointing segmental abnormalities.

This study aims to integrate acoustic signal processing and analysis using AI to develop an innovative diagnostic tool that improves accuracy in respiration analysis and enables easier transition of the analysis of auscultation sounds in the lungs to the digital age. All the gathered data will then be analyzed employing a cocktail of Descriptive Statistical Analysis to gain a sound insight into the acoustic properties of the sounds produced within the lungs depending on the anatomical segments and the study participants. Descriptive statistical analysis will then be performed using mean values with corresponding standard deviations (Mean ± SD) to represent all the demographic factors and the acoustic variables to give a clear indication of the data. Comparisons will be made using Repeated Measures Analysis of Variance (ANOVA) to compare the differences in Root Mean Square (RMS) intensity values depending on the anatomical segments within the study participants to determine the intrapulmonary sound patterns. In cases where there are significant values, the Bonferroni Correction will be employed to correct the Type I error rates occasioned by multiple comparisons to maintain a stringent level of statistical significance at p < 0.005. Further classification of sound intensity levels into low, medium, and high groups using supervised classification modeling with annotated training data derived from the sound data. Statistical analysis will be done using R Studio.

### Harms

The study will include non-invasive auscultation using a validated electronic stethoscope. The potential for harm to participants is to be minimal. Any adverse events such as any discomfort during the process of auscultation, vertigo during breathing, or anxiety during data collection-will be recorded and addressed as soon as possible. The participants will be continuously monitored during the procedure, and data acquisition will be stopped instantly if any signs of distress, shortness of breath, or fatigue appear.

The details would be recorded on an event Record Form regarding the nature of the event, time of onset, duration, severity, and action taken. Serious adverse events, though not expected, would be informed to the Ethics Review Committee, Faculty of Medicine, University of Peradeniya, immediately upon detection within 24 hours. Events and protocol deviations will also be documented in the participant’s CRF with due consideration for transparency and traceability.

### Ethical Considerations

This study has sought approval from the ethics committee, which is in the final approval stage among the Faculty of Medicine, University of Peradeniya, Sri Lanka, as per ERC Reference Number 2025/EC/87. The responses to all queries have been attended to, and the revised copies have been resubmitted for final approval. The participants will be fully briefed in writing about the aim, process, risks, as well as the benefits associated with the study in detail in the participant’s native language to eliminate the misunderstandings of the participants Before starting data collection, every participant will be asked to sign the consent form. To preserve participant privacy, the participants’ identifiers will be replaced by participant code numbers. No identifiers shall be presented in the dataset.

The datasets shall be stored on encrypted servers only accessible by the two chief investigators. Hard copies of the documents would be placed in a locked filing cabinet in secured facilities.

To ensure the full preservation of participant integrity, the raw datasets shall be archived on the servers for five years only after completion. These shall be irreparably erased using industrial-standard software.

## Discussion

Auscultation is the traditional focal point for the assessment of respiration; however, it is inherently subjective. The current existence of digital auscultation technologies fills the gap that was previously present due to the lack of quantitative results. The protocol provides an effective framework that enables the collection, interpretation, and standardization of the analyzed data regarding lung sounds.

Prevailing and already established respiratory sound datasets auscultation datasets including the ICBHI 2017 Respiratory Sound Database, HF Lung V1 database, PERCH digital auscultation datasets, have significantly contributed to the advancement of AI-assisted respiratory diagnostics by enabling the development of machine learning models for wheeze, crackle, and disease classification. The ICBHI 2017 database became a widely used benchmark dataset for respiratory sound classification research and has been extensively utilized for training convolutional neural networks and feature-based classification algorithms.

Most existing repositories primarily focus on disease-level or patient-level classification rather than segmental or lobar acoustic characterization. In many databases, respiratory sounds are recorded from limited chest locations without systematic anatomical standardization, reducing the ability to investigate regional airflow heterogeneity and localized pathological acoustic changes Another limitation of existing respiratory sound datasets is the lack of detailed metadata integration.

The present study aims to address these methodological and geographical gaps by developing a standardized pulmonary sound repository using standard anatomical auscultation landmarks, structured inspiratory-expiratory annotations, and comprehensive demographic and physiological metadata collection. Unlike many existing repositories, the current protocol incorporates both waveform-based and spectrogram-assisted respiratory phase annotation together with region-specific recording methodology across multiple lung segments. The inclusion of lobar-level respiratory recordings may improve future understanding of regional acoustic features in obstructive lung diseases and support the development of more anatomically sensitive AI classification systems.

In addition, the proposed repository is intended to establish a structured and reproducible framework for future respiratory acoustics research in low-resource and telemedicine settings. The use of portable digital stethoscope technology together with standardized preprocessing and annotation protocols may improve dataset reproducibility and facilitate future interoperability with international respiratory sound databases. Ultimately, this repository may contribute toward the development of more robust, regionally adaptable, and clinically interpretable AI-assisted auscultation systems for respiratory disease assessment, With the emphasis on segmental analysis, the current research can actually exceed the traditional global approach to analyzing the distribution of airflow. This study provides more information on the effect of anatomical characteristics on sound transmission, an important consideration for the training of the algorithm for the system.

### Strengths of this study

It aims to examine segmental and lobar differences in lung sound properties among patients with obstructive lung diseases and healthy participants through reliable digital auscultation.

This research aims to develop a quality database of digital lung sounds. This database will be of great use to all professionals involved in the field of respiratory diagnostics and physiotherapy.

The integration of robust acoustic analysis methodologies such as Root Mean Square energy, Mel-Frequency Cepstral Coefficients, and spectrographic mapping with machine learning algorithms is an ongoing innovative effort in the use of AI for the classification and diagnosis of respiratory disease.

The fact that these digital stethoscopes are non-invasive and portable makes them very feasible and scalable in resource-challenged settings.

### Limitations

The study is conducted as a single center study at a tertiary care centre On the plus side, the repository could start with a relatively small set of participants, compared to international studies, which could mean that there is limited diversity of the AI model.

## Author Contributions

Conceptualization: Anuradha RPH, Yasaratne D, Godaliyadda GMRI.

Methodology: Anuradha RPH, Yasaratne D, Ekanayake MP, Siverin R.

Software and analysis: Godaliyadda GMRI, Ekanayake MP.

Writing – original draft: Anuradha RPH.

Writing – review & editing: Yasaratne D.

All authors approved the final manuscript.

## Trial Registration

Not applicable. This study follows SPIRIT 2013 for observational protocols.

## Data Availability

No datasets were generated or analysed during the current study. All relevant data from this study will be made available upon study completion.

